# KASP-PCR method to screen thrombophilia genetic risk factors

**DOI:** 10.1101/2023.10.25.23297518

**Authors:** Reham Altwayan, Huseyin Tombuloglu, Abdulrahman Alhusil, Taghreed Awadh, Mona Altwayan, Heba Albaqawi, Noof Aldossary, Turgay Unver

## Abstract

Thrombophilia is defined as the willingness of blood to clot easily in a situation of imbalances between fibrinolysis and coagulation. It is classified as inherited and acquired thrombophilia. Several studies revealed that the inherited thrombophilia is strongly associated with single nucleotide polymorphisms (SNP) or deletions on certain genes, such as *FV Leiden (rs6025), MTHFR1 (rs1801133), MTHFR2 (rs1801131), Serpine-1 (rs1799768),* and *Factor II (rs1799963)*. This study aims to develop an SNP detection panel based on Kompetitive Allele Specific Primer-polymerase chain reaction (KASP-PCR) technique. Results revealed that 86.5% of susceptible patients (n = 111) contain at least one mutation; while seven of them harbor three thrombophilia-associated mutations simultaneously. A clear allelic discrimination was observed for all tested samples. The prevalence of each mutation among different countries and ethnic groups are in line with the findings of this study. Rather than expensive and time-consuming approaches, the current assay enables the cost-effective advantage of the KASP-PCR, which reduces the diagnostic cost with a fast and convenient way. After clinical validation and approval, it can be used in hospitals, research centers, and diagnostic laboratories to determine the genetic susceptibility of individuals to thrombosis and for research purposes.

## Introduction

Thrombophilia is defined by the World Health Organization/International Society of Thrombosis and Hemostasis (WHO/ISTH) as an abnormal tendency to coagulation or clotting of the blood in a part of the circulatory system (WHO, 1995). The disease poses a significant health burden. According to the CDC reports, >900,000 people in the United States alone are affected by blood clots each year, with substantial mortality rates. Due to thrombophilia-related complications, 60,000 to 100,000 deaths are recorded in the USA annually, which is greater than the total number of people who lose their lives each year from AIDS, breast cancer, and motor vehicle crashes combined (Badireddy & Mudipalli, 2022). The disease is classified as inherited and acquired thrombophilia. Inherited thrombophilia is associated with four SNPs and one deletion mutations located in different genes, namely: coagulation *factor II (FII)*, *coagulation factor V (FV)*, *5,10-methylene tetrahydrofolate reductase (MTHFR1* and *MTHFR2)*, and *serpine1 (SERPINE1)* (Yapijakis et al, 2015).

Diagnostic approaches include detecting disorders by examining clinical signs, imaging techniques, and analysis through molecular aspects. Direct DNA genotyping, PCR-based techniques, immunological detection, and chromogenic assays are used to identify thrombophilia-related disorders (Linnemann and Hart 2019). Sequencing techniques are the most accurate approaches followed and used for validation purposes. On the other hand, PCR-based approaches such as TaqMan, KASP and rhAmp are mostly used due to their simplicity, timesaving, cost-effectiveness, and ability to measure the heterozygosity or homozygosity of mutations carried among thrombophilia genes.

Kompetitive Specific Allele Primer-PCR (KASP-PCR) is a fluorescent-based reporting system to identify and measure the SNPs (He et al., 2014). It is used to differentiate between species in plants and animals (Yang et al., 2020). KASP-PCR technique is fast, sensitive, relevant, and cost-effective (Alvarez-Fernandez et al., 2021; Zhang et al., 2020). Due to the cost advantage, it has been applied widely in plant breeding for commercial use (Makhoul et al., 2020). KASP technique is precise, and its specificity is based on the competitive nature of allele specific forward primers and modified Taq polymerase.

The current genotyping techniques are based on sequencing or PCR, which are time consuming and expensive methods. Therefore, a cost-effective, time-saving, and accurate technique is needed to screen symptomatic and asymptomatic suspicious people. This study aims to screen thrombophilia associated SNPs based on KASP-PCR technique. For this purpose, the DNA samples of thrombophilia-susceptible patients (n = 111) were collected and five associated mutations on *FV Leiden, MTHFR1, MTHFR2, Serpine-1,* and *Factor II* genes were identified. Rather than expensive and time-consuming approaches, the current assay reduces the diagnostic cost in a fast and convenient way.

## Material and Methods

### Collection of blood samples and ethical concerns

5 mL of leftover whole blood samples were retrieved from King Fahad Military Medical Complex (KFMHC) (IRB Protocol No: AFHER-IRB-2022-031). A total of 111 samples have been collected from thrombophilia-susceptible patients who have been admitted to the hospital with thrombophilia symptoms, such as chest pain, abdominal pain, dizziness, fever, seizure, breath shortness, etc. The blood samples were collected in an Ethylenediamine tetra acetic acid tube (EDTA) and stored at −20 °C until DNA extraction.

### Extraction of DNA

QIAamp® DNA Blood Mini Kit 250 (QIAGEN) was used to extract and purify DNA. The instructions of the manufacturer were followed. The final eluate was kept at −20 °C for further stages. NanoDrop^TM^ 2000/2000c spectrophotometer was used to quantify the purity and concentration of extracted DNA samples. The samples were also visualized in agarose gel for 20 min at 100 Volt and monitored under UV light (Molecular Imager® ChemiDoc™ XRS+, Bio-Rad). Image Lab™ Software (Bio-Rad) was utilized to save the images.

### Primer design

Primers corresponding to the SNP positions were manually designed. First, the corresponding gene sequences were retrieved from NCBI database (https://www.ncbi.nlm.nih.gov) and Primer Blast tool was used to pick the primers. In addition, Primer 3 program was used to verify the designed primers: one common reverse, two alternating forward primers that are specific to the SNP positions.

### Validation of plate reader and KASP-PCR reaction

Before genotyping, KASP genotyping validation kit (LOW ROX KASP TF Validation kit, LGC) was used to validate the plate-reader instrument (7500 Fast Real-Time PCR System). According to the manufacturer’s protocol, the compatibility and readability of the plate reader was confirmed. The kit consists of three tubes of diluted fluorophores: FAM, HEX, and HEX/FAM. The tubes were briefly vortexed and dispensed into a microtiter plate. Then, KASP reagents (2x KASP master mix, and 72x KASP assay mix) with provided DNA samples (n = 36) were used to run the following protocol: (1) 94 °C 15 minutes for activation (1 cycle); (2) 94 °C 20 sec for denaturation and 61-55 °C 60 sec (drop 0.6 °C per cycle) for annealing/elongation (10 cycles); (3) 94 °C 20 sec for denaturation and 55 °C 60 sec for annealing/elongation (26 cycles). At the end of the reaction, the endpoint fluorescent reading was performed. To obtain the best genotyping clusters, the plate underwent two rounds of recycle steps, as follows: 94 °C 20 sec for denaturation, and 57 °C for 60 sec for annealing/elongation (3 cycles).

### Genotyping of thrombophilia-associated SNPs with KASP-PCR

The KASP primer mixture for each target SNPs was prepared including 12 µl of forward-1 primer, 12 µl of forward-2 primer, 30 µl of common reverse primer, and 46 µl of distilled water to reach the total volume of 100 µl. Then, 5 µl of master mixture (KASP™ TF Low ROX Master Mix) and 0.14 µl of assay mixture were added and vortexed briefly. In 96 well plate, 5 µl of assay mixture was added with 5 µl of DNA sample in each well and then sealed tightly. The KASP genotyping assay was performed by using real-time PCR 7500 (Applied Biosystem) instrument.

## RESULTS

### Calibration and optimization of KASP protocol

Before the KASP-PCR reaction for the discrimination of thrombophilia-associated SNPs, the validity of the fluorescent thermal cycler and the protocol for KASP-PCR were optimized and validated. Nine positive control samples (LGC Bioscience), labeled with FAM (blue), HEX (red), and FAM/HEX (green) fluorophores, were used to optimize, and verify the fluorescence signals that will be obtained from the thermal cycler. **Figure 1** depicts the cluster plot of FAM, HEX, and FAM/HEX labeled positive control samples. The clusters correspond to homozygote dominant (allele 1/allele 1), homozygote recessive (allele 2/allele 2), and heterozygote (allele 1/allele 2) discriminations. This result showed optimized and successful KASP-PCR reaction conditions with calibrated fluorometric measurements.

**Figure 1.**
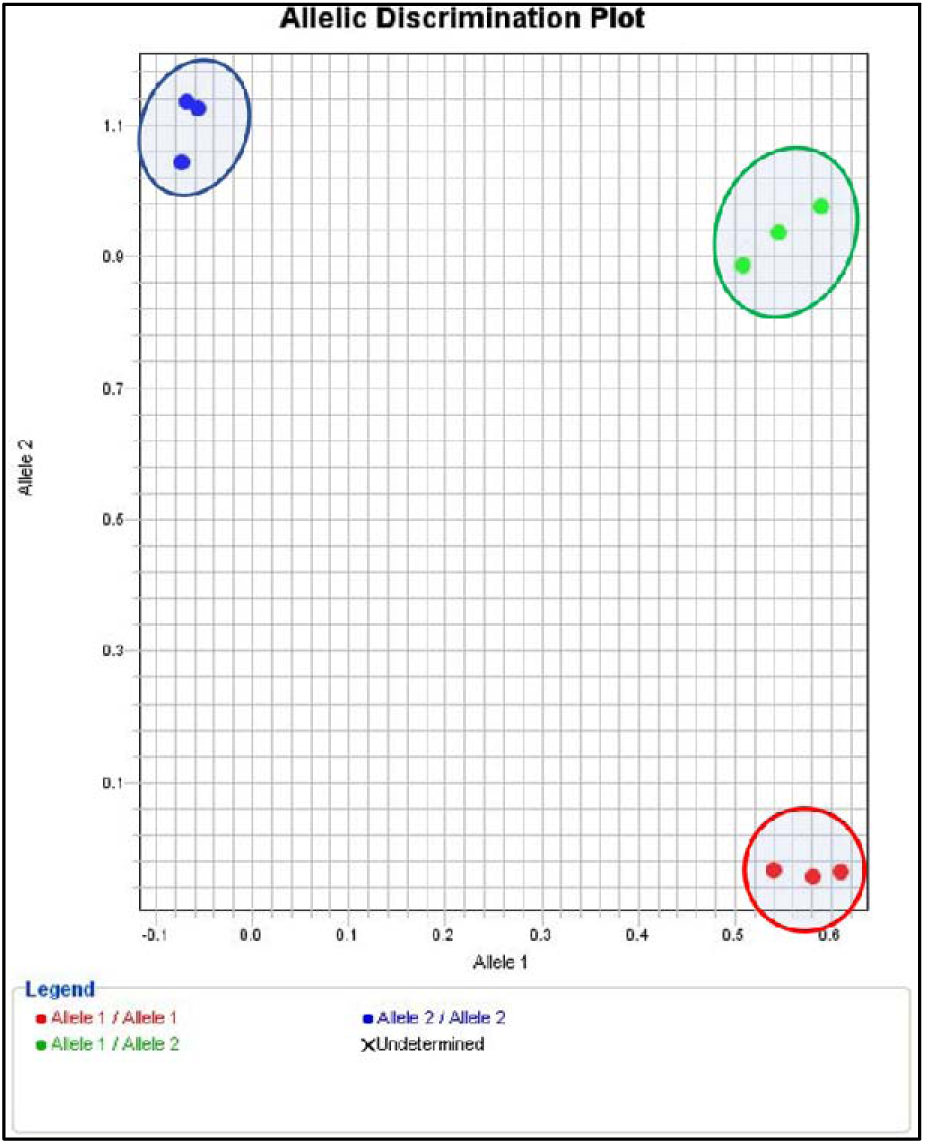
The allelic discrimination plot of FAM (blue), HEX (red) and FAM/HEX (green) fluorophores. Three samples used for each positive control dyes.

Furthermore, the optimized protocol was used to discriminate real DNA samples (n = 45) provided by the LOW ROX KASP TF Validation kit (LGC Biosciences). In addition to the DNA samples, three negative control samples were included. After the KASP-PCR reaction, the allelic discrimination plot was depicted in **Figure 2**. The results were shown before (**Figure2A**) and after three (**Figure2B**) and six **(Figure2C**) additional recycling stages. After six recycling stages, positive and negative samples were clearly separated, and the negative control specimens were placed near the origin side of the plot. In addition, the homozygote and heterozygote alleles were determined to be well-clustered. It is obvious that additional recycling steps improved the allelic discrimination, where the best and most efficient results were obtained upon six additional recycling stages (**Figure 2C**).

**Figure 2.**
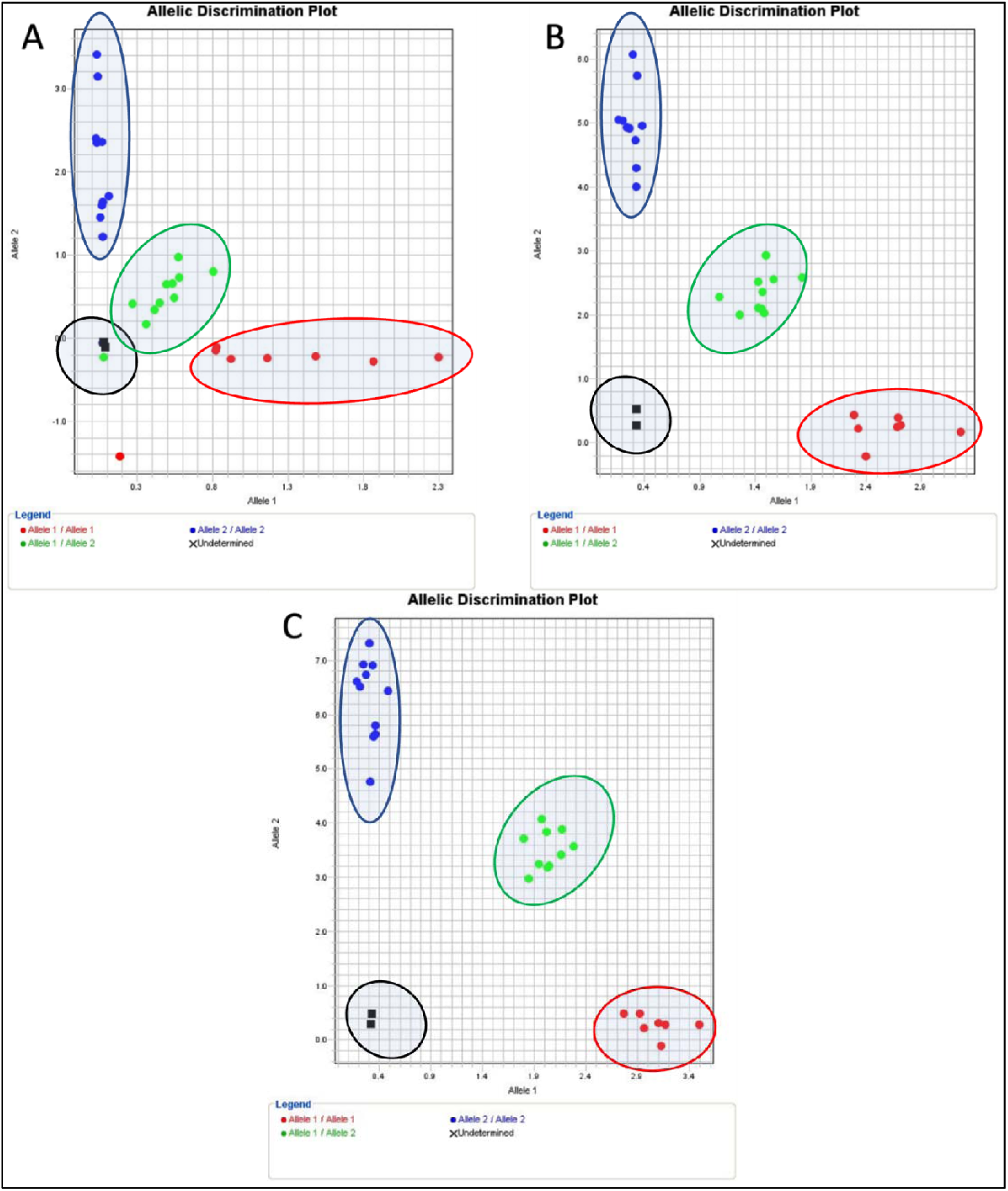
Validation of plate reader and KASP-PCR protocol. Allelic discrimination plot of DNA samples (n = 45). A) Without recycle step. B) After the addition of three recycling steps. C) After the addition of six recycling steps. Black dots indicate the negative control sample. Red, blue, and green dots indicate the homozygotes of Allele 1, homozygotes of Allele 2, and heterozygotes of Allele 1/2.

### KASP-PCR genotyping of rs6025 (G1691A) on the Factor V Leiden gene

An optimized KASP-PCR protocol was performed to identify the G1691A substitution on the *FVL* mutation (*FV* gene, rs6025). **Figure 3**. shows the discrimination plots of samples. Since the total sample number (n = 111) is more than the capacity of the thermal cycler (n = 96), the reaction was completed in two separate runs. Upon the KASP-PCR reaction, an additional six recycling steps was added for the best reaction performance. Black dots indicate negative control, green dots indicate heterozygote sample (CT), and blue dots indicate homozygote (CC). Accordingly, only one sample was detected as heterozygote (∼1%), and the rest of the samples were homozygote wild (∼99%). This result shows that almost all thrombophilia-susceptible individuals admitted to the hospital are free of *FVL* gene mutation.

**Figure 3.**
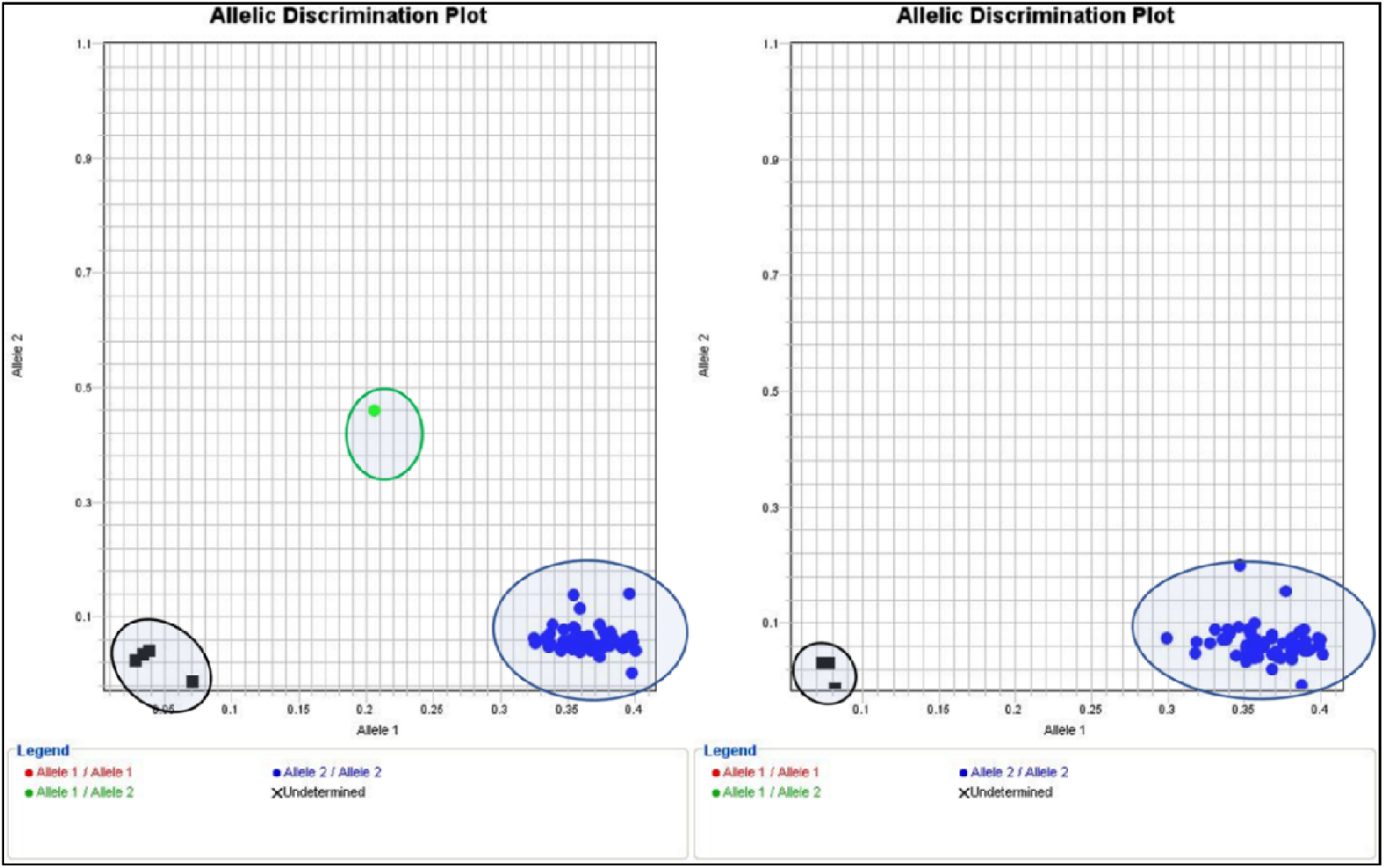
The figure shows the discrimination plot of FVL mutation with an additional 6 cycles. Black dots indicate negative control, green dot indicate heterozygote sample (CT), and blue dots indicate homozygote wild (CC).

### KASP-PCR genotyping of (rs1799963) (G20210A) on the *FII* gene

KASP-PCR assay was performed to screen the G20210A variation on the *FII-PT* gene. **Figure 4.*1*** depicts the discrimination plots of extracted DNA samples. Since the capacity of the PCR plate is 96 wells, the run was divided into two runs. In addition to the KASP-PCR thermal cycle, six more recycling step was added to improve discrimination. Black dots refer to negative control, placed near the plot’s origin side; blue dots refer to homozygote wild samples (GG); red dot sample is homozygote mutant (AA), while no green dots of heterozygote recorded. Only one sample was recorded as homozygote recessive, while the rest of the sample was homozygote dominant, and no heterozygote detected. This result shows that only one of the susceptible thrombophilia patients carried the *FII-PT* mutation.

**Figure 4.1.**
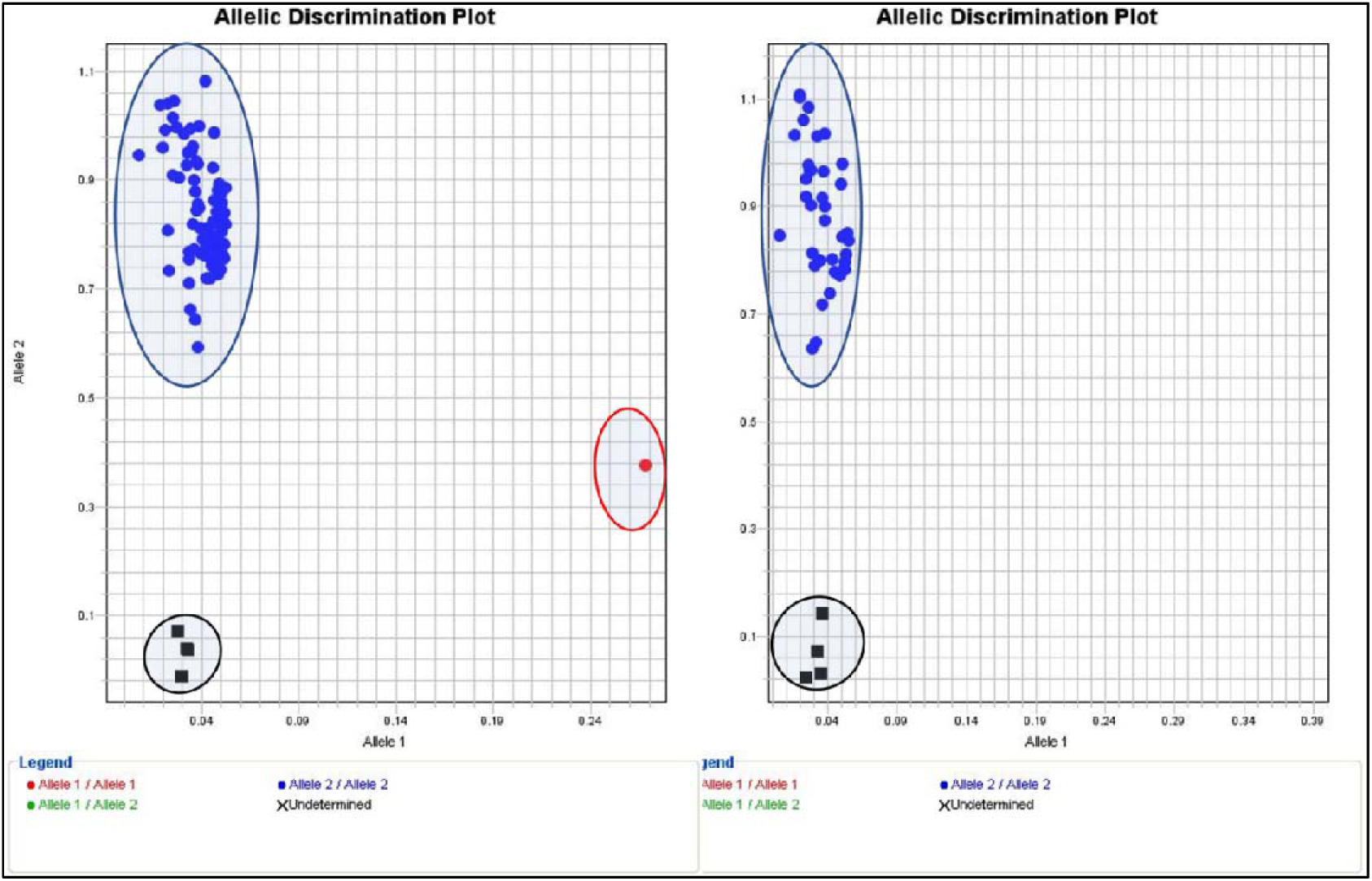
Shows the discrimination plot of *Factor II mutation* with an additional 6 cycles. Black dots indicate negative control, blue dots indicate homozygote wild (GG), and red dots indicate homozygote mutant (AA).

### KASP-PCR genotyping of (rs1801133) on the *MTHFR* gene

After optimization of the KASP-PCR assay, screening for mutation on *MTHFR1* was performed (*MTHFR* gene, rs1801133). **Figure *2*5** illustrates the discrimination plot of susceptible-thrombophilia patient samples. The reaction was completed in two runs due to the number of samples (n=111). In addition, six recycling steps were added to obtain clear discrimination. Four black dots refer to negative control and are correctly placed near the origin of the plot, green dots refer to the heterozygote sample (CT), and red dots refer to the homozygote sample (CC). Consequently, 24 % of the patients were heterozygote, only two samples were recorded as homozygote recessive (TT) (∼2%), and the rest of the samples were recorded as homozygote dominant. This result shows that ∼26% of the patients admitted to the hospital with a risk of developing thrombophilia are carrying at least one allele of *MTHFR1* mutation.

### KASP-PCR genotyping of (rs1801131) on the *MTHFR2* gene

An optimized KASP-PCR assay was achieved to screen the mutation placed on the *MTHFR2* gene (rs1801131). **Figure 3** demonstrates the discrimination plot of two runs performed on extracted DNA samples. With the addition of six thermal recycling steps, the reaction shows the best performance. Black dots imply negative control; green dots imply heterozygote sample (AC), blue dots imply homozygote wild (AA), and red dots imply homozygote recessive (CC) alleles. Discrimination results showed that ∼53% of the patients admitted to the hospital are free of mutation (wild), 39% carrying this mutation in one allele, and 6.3% carrying two alleles of mutation. The result shows that ∼45% of susceptible thrombophilia patients have at least one mutation.

**Figure 2.**
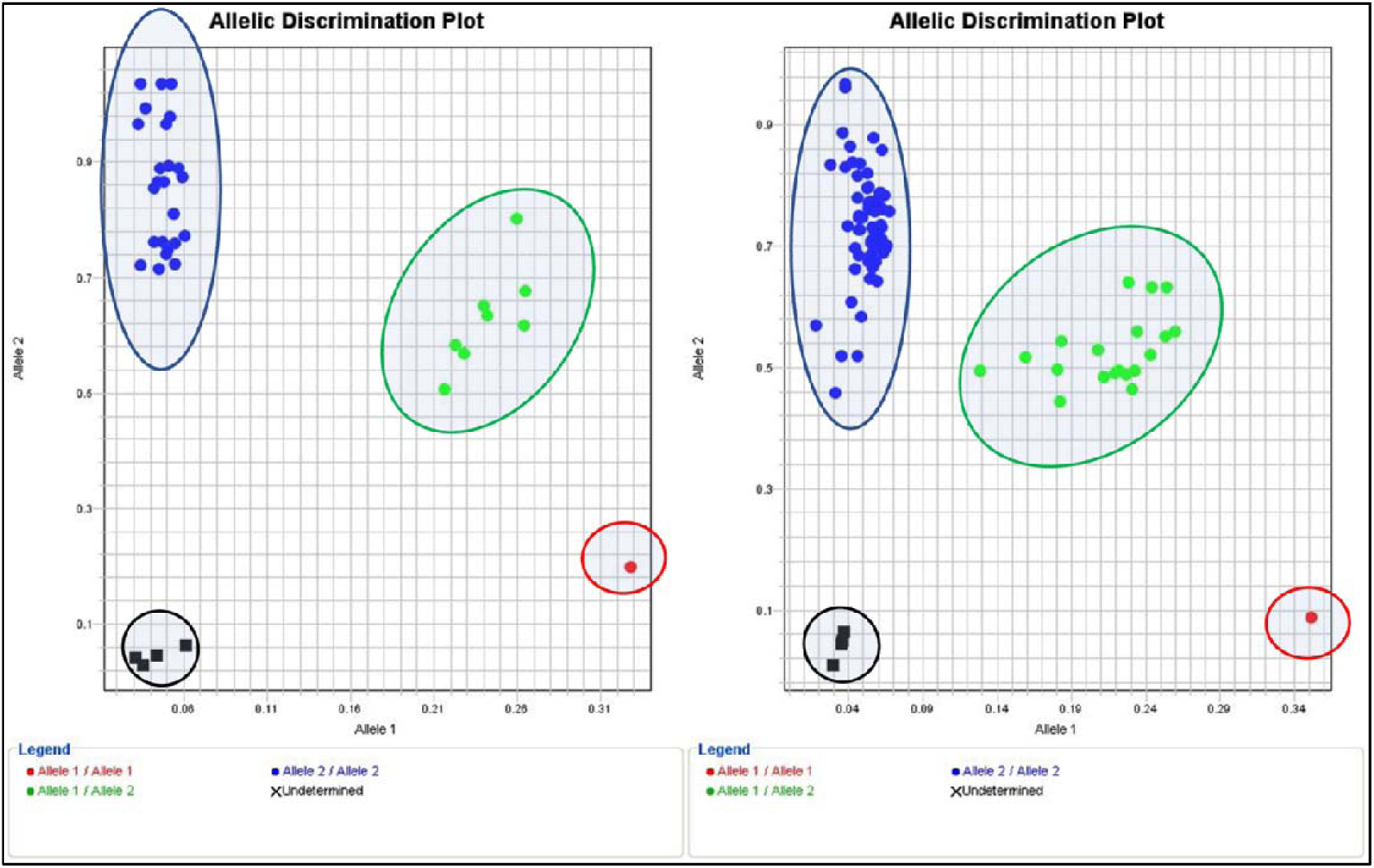
Shows the Allelic Discrimination Plot of *MTHFR* rs1801133 (C/T) mutation with an additional 6 cycles. Black dots indicate negative control; green dots indicate heterozygote (CT); and red dots indicate homozygote mutant (TT); blue dots indicate homozygote.

**Figure 3.**
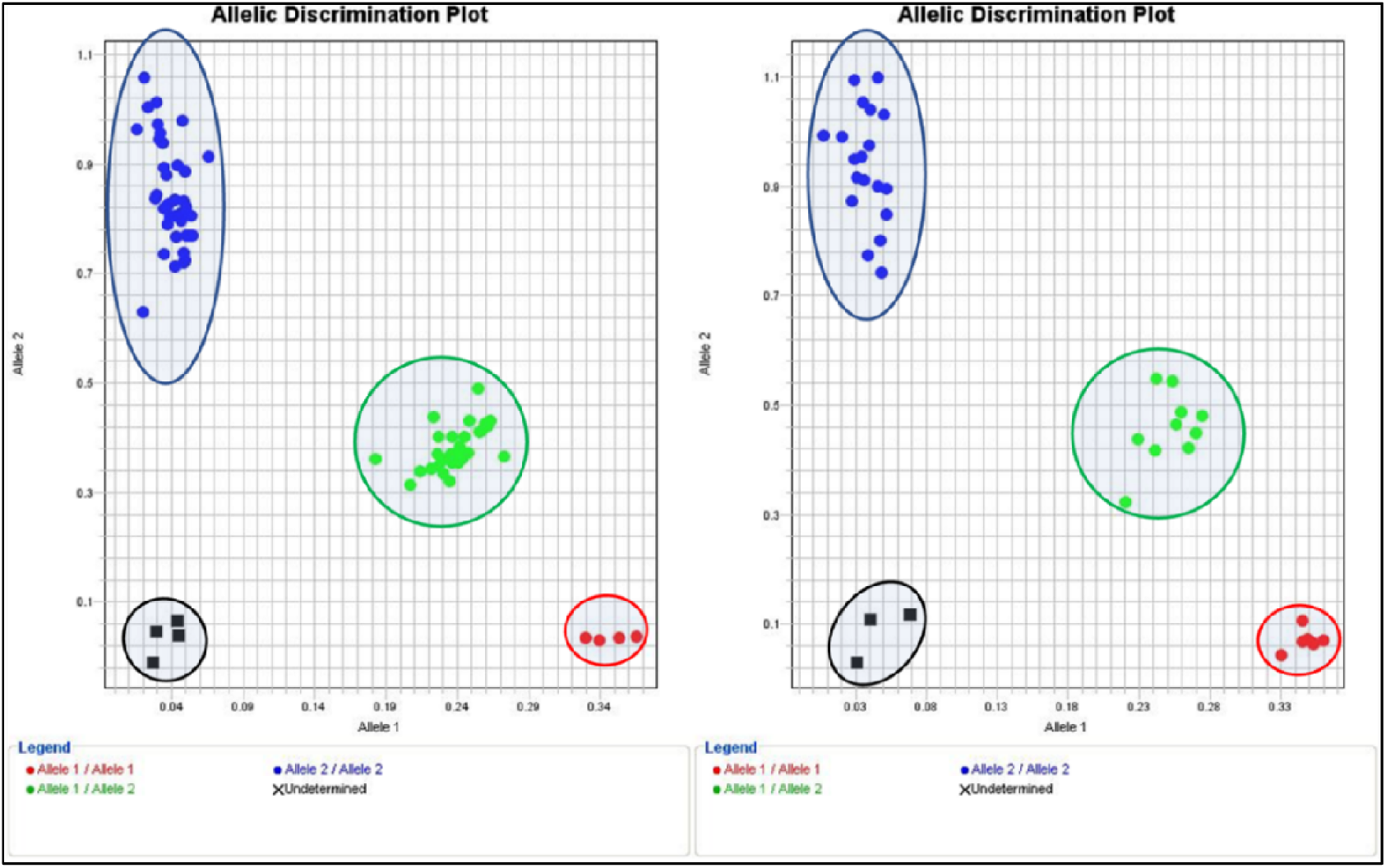
Shows the Allelic Discrimination Plot of *MTHFR* rs1801131 mutation with the addition of 3 cycles. Black dots indicate negative control; red dots indicate homozygote mutant (CC), green dots indicate heterozygote (AC), and blue dots indicate homozygote wild.

### KASP-PCR genotyping of *Serpine 1* (rs1799768)

Genetic screening for *Serpine 1* (rs1799768) mutation was performed by optimized KASP-PCR assay. **Figure 7** shows the discrimination plot of the (n=111) sample divided into two runs due to the capacity of the thermal cycler. Black dots indicate negative control, and it is successfully placed near the origin of the plot; blue dots indicate homozygote dominant 5G/5G, green dots indicate heterozygote 5G/4G and red dots indicate homozygote recessive 4G/4G. Unlike other genes, only three additional thermals are needed to obtain clear discrimination of samples. Therefore, 47 of the sample was detected as homozygote dominant (∼42%), 44 of the sample were detected as heterozygote by carrying one mutation (∼39%), and 17 were detected as homozygote recessive (∼15%). The result shows that more than 50% of patients harbor *Serpine 1* mutation, which increases the risk of developing thrombophilia.

**Figure 7.**
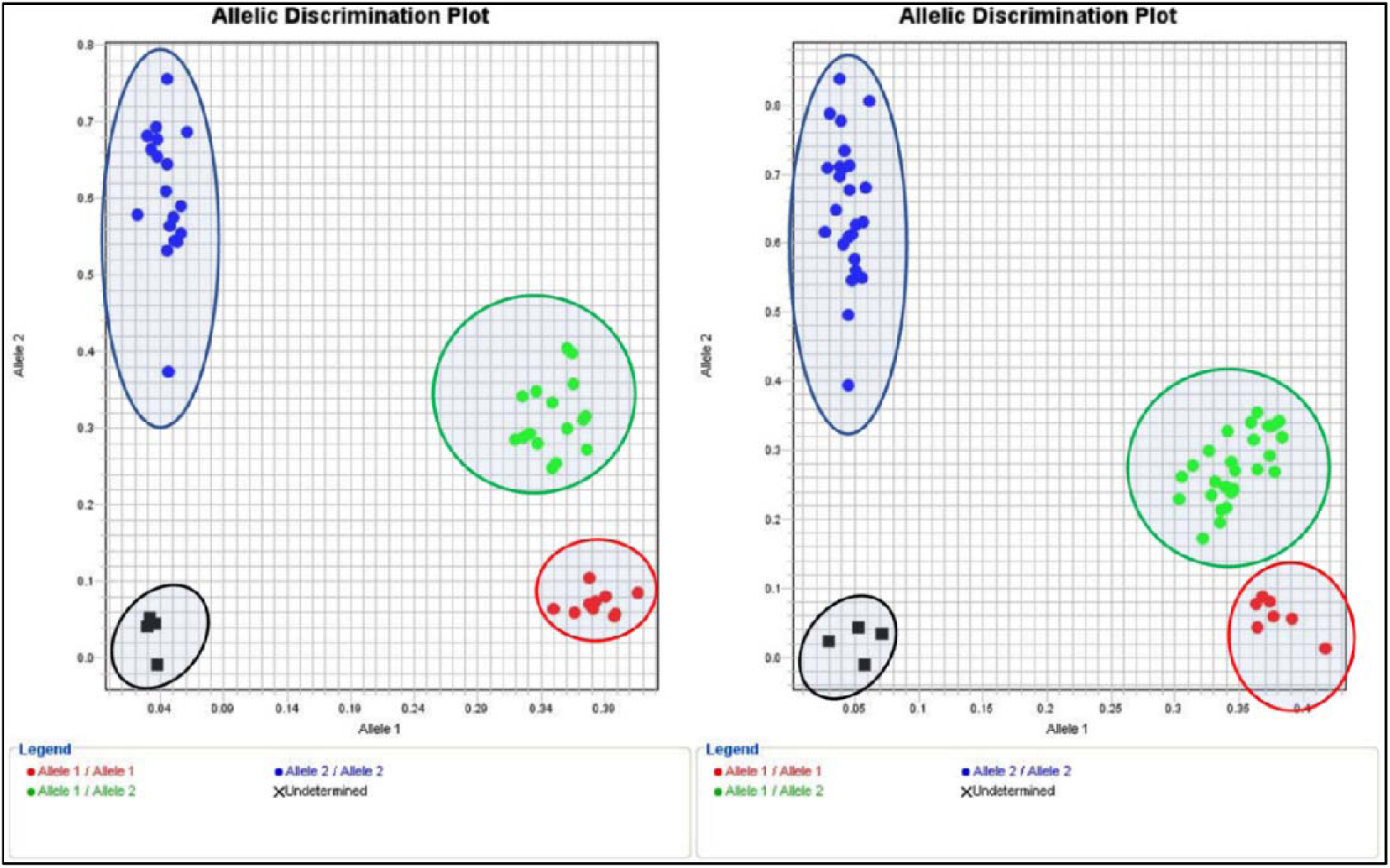
Shows the discrimination plot of *Serpine* 1 rs1799768 mutation with an additional 3 cycles. Black dots indicate negative control, red dots indicate homozygote mutant (4G/4G), green dots indicate heterozygote (5G/4G), and blue dots indicate homozygote wild.

## DISCUSSION

This study aims to screen genetic risk factors in patients susceptible to thrombophilia disease by applying the cost-effective PCR technique (KASP-PCR). For this purpose, left-over blood samples (n = 111) collected from thrombophilia symptomatic patients were utilized for genetic screening. Approximately 30% of the cases were admitted to the hospital with chest pain and 20% with abdominal pain. In addition to these symptoms, vomiting (2.7%), shortness of breath (11%), palpitations (4.5%), dizziness (10%), and fever (5.5%) were among the most common symptoms recorded. Patients who apply to the hospital with suspicion of the disease are aged 5-92 (mean = 52), male and female (62 male, 50 female).

KASP-PCR analyses revealed that 92 out of 111 (∼83%) patients susceptible to thrombophilia carry at least one of the analyzed mutations. The age and gender were not found to be associated with thrombophilia-related polymorphisms (*P* >0.05). Before KASP-PCR genotyping testing, the KASP genotyping validation kit was used to optimize and validate the plate reader. The device was calibrated according to the passive reference dye ROX™ contained in the KASP master mixture. Optimal FAM, HEX, and FAM/HEX fluorescence signals were checked and adjusted for the most efficient signal score. **Figure 1**. shows successful discrimination in testing FAM, HEX, and FAM/HEX dyes; where FAM is clustered in the Y-axis direction, HEX is clustered in the X-axis opposite direction, while the FAM/HEX mixture is clustered in the middle. Moreover, the cycle and re-cycling stages were optimized to obtain the best allelic discrimination (**Figure 2**). Results revealed that two rounds of three re-cycling steps (six in total) provided the most efficient protocol for robust allelic discrimination.

The primary role of the *FV* gene is regulating the production of coagulation factor V protein. A mutation found in this gene (G1691A) substitutes arginine to glutamine amino acid at point 506 in a polypeptide of Factor V (Arg506Gln) (Madkhaly et al., 2021), leading to unresponsiveness to anticoagulant factors (C and S) (Kujovich, 2018). The risk of developing thrombophilia is five times higher in heterozygotes and 50 times higher in homozygotes, and it is responsible for 20-25% of all VTE cases (Bezgin et al., 2018). It was found to be the most critical genetic risk factor related to thrombophilia (Djordjevic et al., 2012). The prevalence of *FVL* among the European population is high, while it is rare in Eastern Asia and Africa. Since the Arab countries are geographically at the center. The prevalence of *FVL* mutation in different countries and ethnic groups are listed in **Table 1**. Our results showed that all collected samples are homozygote wild, except one sample which is heterozygote (No. 61). In **Figure 3**., the heterozygote individual was admitted to the hospital with flank pain and dysuria. A prospective study aims to evaluate the prevalence of *FVL* in 149 healthy Saudi subjects revealed that 2% of the individuals are heterozygotes, and none of those individuals are homozygotes mutant (0%) (Almawi et al., 2005). Golestani et al., (2022) investigated the correlation between acute myocardial infarction (AMI) and *FVL* in the Iranian population; 5.5% of cases were found to be heterozygotes, while 1% were homozygote mutants. On the other hand, Gowda et al., (2000) found that among American subjects, 8% were heterozygote. Based on these results, the data in the literature is in line with and compatible with our results.

**Table 1.**
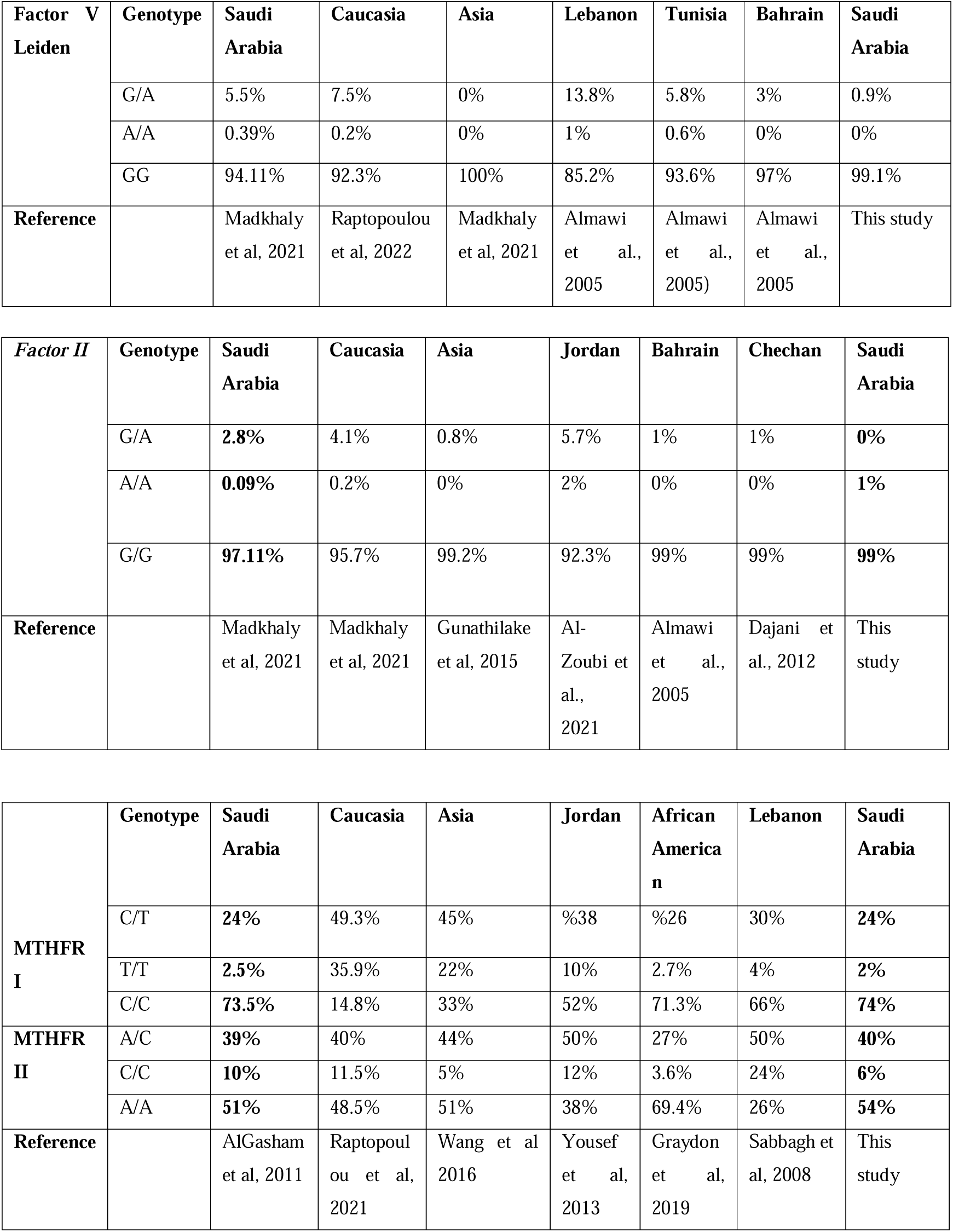
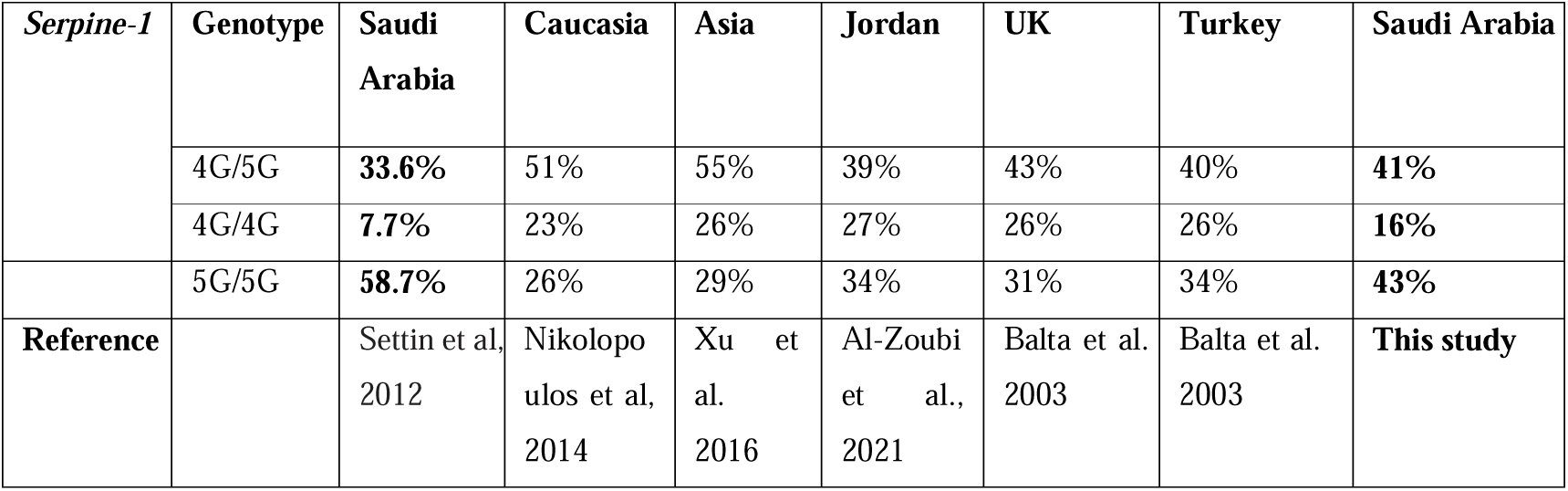
The prevalence of thrombophilia-associated SNPs according to different countries, ethnic groups, and this study.

| **Factor V**<br>**Leiden** | **Genotype** | **Saudi**<br>**Arabia** | **Caucasia** | **Asia** | **Lebanon** | **Tunisia** | **Bahrain** | **Saudi**<br>**Arabia** |
| --- | --- | --- | --- | --- | --- | --- | --- | --- |
|  | G/A | 5.5% | 7.5% | 0% | 13.8% | 5.8% | 3% | 0.9% |
|  | A/A | 0.39% | 0.2% | 0% | 1% | 0.6% | 0% | 0% |
|  | GG | 94.11% | 92.3% | 100% | 85.2% | 93.6% | 97% | 99.1% |
| **Reference** |  | Madkhaly<br>et al, 2021 | Raptopoulou<br>et al, 2022 | Madkhaly<br>et al, 2021 | Almawi<br>et al.,<br>2005 | Almawi<br>et al.,<br>2005) | Almawi<br>et al.,<br>2005 | This study |

| **Factor II** | **Genotype** | **Saudi**<br>**Arabia** | **Caucasia** | **Asia** | **Jordan** | **Bahrain** | **Chechan** | **Saudi**<br>**Arabia** |
| --- | --- | --- | --- | --- | --- | --- | --- | --- |
|  | G/A | **2.8%** | 4.1% | 0.8% | 5.7% | 1% | 1% | **0%** |
|  | A/A | **0.09%** | 0.2% | 0% | 2% | 0% | 0% | **1%** |
|  | G/G | **97.11%** | 95.7% | 99.2% | 92.3% | 99% | 99% | **99%** |
| **Reference** |  | Madkhaly<br>et al, 2021 | Madkhaly<br>et al, 2021 | Gunathilake<br>et al, 2015 | Al-Zoubi et<br>al.,<br>2021 | Almawi<br>et al.,<br>2005 | Dajani et<br>al., 2012 | This<br>study |

|  | **Genotype** | **Saudi**<br>**Arabia** | **Caucasia** | **Asia** | **Jordan** | **African**<br>**America**<br>**n** | **Lebanon** | **Saudi**<br>**Arabia** |
| --- | --- | --- | --- | --- | --- | --- | --- | --- |
| **MTHFR**<br>**I** | C/T | **24%** | 49.3% | 45% | %38 | %26 | 30% | **24%** |
|  | T/T | **2.5%** | 35.9% | 22% | 10% | 2.7% | 4% | **2%** |
|  | C/C | **73.5%** | 14.8% | 33% | 52% | 71.3% | 66% | **74%** |
| **MTHFR**<br>**II** | A/C | **39%** | 40% | 44% | 50% | 27% | 50% | **40%** |
|  | C/C | **10%** | 11.5% | 5% | 12% | 3.6% | 24% | **6%** |
|  | A/A | **51%** | 48.5% | 51% | 38% | 69.4% | 26% | **54%** |
| **Reference** |  | AlGasham<br>et al, 2011 | Raptopoulou<br>et al, 2021 | Wang et al<br>2016 | Yousef<br>et al,<br>2013 | Graydon<br>et al,<br>2019 | Sabbagh et<br>al, 2008 | This<br>study |

| *Serpine-1* | Genotype | Saudi Arabia | Caucasia | Asia | Jordan | UK | Turkey | Saudi Arabia |
| --- | --- | --- | --- | --- | --- | --- | --- | --- |
|  | 4G/5G | **33.6%** | 51% | 55% | 39% | 43% | 40% | **41%** |
|  | 4G/4G | **7.7%** | 23% | 26% | 27% | 26% | 26% | **16%** |
|  | 5G/5G | **58.7%** | 26% | 29% | 34% | 31% | 34% | **43%** |
| **Reference** |  | Settin et al, 2012 | Nikolopoulos et al, 2014 | Xu et al, 2016 | Al-Zoubi et al., 2021 | Balta et al. 2003 | Balta et al. 2003 | **This study** |

*Prothrombin* is a protein synthesized in the liver and is essential in clotting the blood (Khan & Dickerman, 2006). As a result of the replacement of guanine with adenine at position 20210 of the *coagulation factor II (FII)* gene (G20210A), high concentrations of prothrombin protein are activated (Poudel et al., 2020; Ahmed et al., 2020; Rennert & DeSimone, 2019). An increase in prothrombin level increases thrombin level and inhibits Activated Protein C (APC) - inhibits FV protein-, subsequently stimulating thrombus formation. Thus, the risk of VTE appearance increases. It is estimated to be the second most important genetic risk factor causing thrombophilia (Djordjevic et al., 2012). In our experiment, one out of 90 patients carry a homozygous mutant allele (Patient No. 52, 86 years old male, and suffering from dysuria); no mutation was detected in other samples (**Figure 4.*1*4**). A study has been conducted to screen *FII* and *FVL* mutations from eight years-old patient who is suffering from dysuria and priapism. The genotyping result revealed that the patient was heterozygote for both mutations (Özbek et al., 2003). At the same time, the patient is homozygote with *FII* and heterozygote with *FVL*. A retrospective study showed that 2.6% of patients screened for *FII* are positive through Saudi thrombosis patients (Madkhaly et al., 2021). A cross sectional study investigate the prevalence of *FII* mutation in Jordanian population; 5.7% were heterozygote while 2% were homozygote mutant (Al-Zoubi et al., 2021). A group of healthy Arab population enrolled in a retrospective study to test the distribution of *FII*. No mutated allele among Saudis was observed, while 1% of heterozygotes in Bahraini. In Lebanese and Tunisian, 3.6% and 2.6% are heterozygotes, respectively (Almawi et al., 2005). An amplification refractory mutation detection system (ARMS) was applied to define the prevalence of *FII* in Chechans populations in Jordan; only 2% of heterozygotes were detected, while no homozygote mutant (Dajani et al., 2012). The fact that the mutation prevalence in the general population is 2-3% in the western region, while rare in other geographical regions and all previous literature shows a rare incidence of FII mutation, which reflects KASP-PCR results.

The *MTHFR* gene decreases homocysteine concentration by producing the MTHFR enzyme to convert the homocysteine to methionine (Hickey et al., 2013; Dean, 2016). Two recognized mutations in the *MTHFR* gene inactivate the enzyme, leading to increased levels of homocysteine and decreased levels of folate in plasma: C677T (rs1801133) and A1286C (rs1801131) (Dölek et al., 2007). The American Congress of Obstetricians and Gynecologists recommends screening for *MTHFR* mutations instead of homocysteine concentration in patients at risk of thrombophilia (Hickey et al., 2013). In our experiment, we detected these mutations using KASP-PCR method. The prevalence of C677T (rs1801133) was ∼2% with homozygote mutant, ∼24% heterozygote, and ∼75% homozygote wild (**Figure *2*5**). Whereas the prevalence of A1286C (rs1801131) was ∼6% homozygote, ∼40% heterozygote, and ∼55% homozygote wild (**Figure *3*6**). The prevalence of C677T among Hispanics is over 25% and between 10-15% among Caucasians and North Americans with homozygote type (Hickey et al., 2013). Dajani et al, (2013). Investigating the prevalence of C677T among Chechen and Circassians healthy individuals; 27.5% of Chechen carry the mutation, while 50% of Circassians. A study reported that 31% of patients with stroke carried heterozygosity of C677T, while 15% were homozygote mutants. The higher incidence of that variant is attributed to other risk factors, such as smoking and hypertension (Djordjevic et al., 2012).

A variation in the *serpine 1* gene is represented by the deletion of 4G/5G alleles in position 675 in chromosome 7 at the promoter region (Xu et al., 2016; Isordia-Salas et al., 2009; Tsai et al., 2008). The role of the *serpine1* gene is to synthesize plasminogen activator inhibitor 1 (PAI-1), and the mutation increases production (Morange et al., 2007). High concentration of PAI-1 leads to inhibit plasmin from fibrinolysis, causing to accumulate clots in veins (Heit MD, 2013; Morange et al., 2007; Kazuo Miyashita et al., 2012). The allelic discrimination of KASP-PCR of this mutation showed that ∼40% of samples are heterozygote, while 14.4% were detected as homozygote mutant (**Figure 7. 7**). In total, 54% of patients carried PAI-1 variation. Yapikajis et al., (2012) conducted a genotyping study using blood samples from a healthy Greek population, and 49% were found mutated. A case-control study was performed to define the correlation between high concentration of PAI-1 and developing ischemic stroke in young Indian patients. Akhter et al., (2017) found that 62% of young Indian patients have PAI-1 SNP. Xu et al., (2016) found that ∼46% of chronic obstructive pulmonary disease through Chinese Han patients were heterozygote, while ∼14% were homozygote mutants. These results are in line with the current findings (Table 1).

## Conclusion

A gene panel covering five thrombophilia associated mutations (*Serpine 1, MTHFR1, MTHFR2, Factor V Leiden, and Factor II*) was developed based on the KASP-PCR assay to diagnose thrombophilia genetic risk factors. The variety of diagnostic tests, the unreliability of clinical judgment, and a delayed diagnosis could lead to illnesses that may increase mortality and morbidity. In addition, molecular-based analyses are highly costly and time-consuming to obtain accurate results compared to the KASP-PCR assay. The KASP-PCR assay is a cost-effective (∼2-3 dollar per reaction), timesaving (3-4 hours), non-invasive, accurate, and highly sensitive. In this study, we obtained a clear allelic discrimination for all samples tested to screen SNPs related to thrombophilia. Rather than expensive and time-consuming approaches, the current assay enables the cost-effective advantage of the KASP-PCR assay, which reduces the diagnostic cost with a fast and convenient way. After clinical validation and approval, it can be used in hospitals, research centers, and diagnostic laboratories to determine the genetic susceptibility of individuals to thrombosis and also for research purposes.

## Data Availability

All data produced in the present study are available upon reasonable request to the authors

## Acknowledgment

This study is funded by the Deanship of Graduate Studies under project number 2023-003-IRMC. The figures are created with BioRender.com.

## Compliance with Ethical Standards

## Authors’ contributions

HT and RA conceptualized the study. RA and HT conducted the experiments. RA, and HT wrote the manuscript. AA, TA, MA, HA, NA, AIA identified the patients and provided the samples. TU revised and commented on the manuscript.

## References

Wang X, Fu J, Li Q, Zeng D. Geographical and Ethnic Distributions of the MTHFR C677T, A1298C and MTRR A66G Gene Polymorphisms in Chinese Populations: A Meta-Analysis. PLoS One. 2016 Apr 18;11(4):e0152414. doi: 10.1371/journal.pone.0152414. PMID: 27089387; PMCID: PMC4835080.

Akhter MS, Biswas A, Abdullah SM, Behari M, Saxena R. The Role of PAI-1 4G/5G Promoter Polymorphism and Its Levels in the Development of Ischemic Stroke in Young Indian Population. Clin Appl Thromb Hemost. 2017 Nov;23(8):1071–1076. doi: 10.1177/1076029617705728. Epub 2017 May 1. PMID: 28460568.

Algasham, A., Ismail, H., Dowaidar, M., & Settin, A. A. (2011). Methylenetetrahydrofolate Reductase (MTHFR) and Angiotensin Converting Enzyme (ACE) Gene Polymorphisms among Saudi Population from Qassim Region. International Journal of Health Sciences, 5(2 Suppl 1), 3.

Almawi WY, Keleshian SH, Borgi L, Fawaz NA, Abboud N, Mtiraoui N, Mahjoub T. Varied prevalence of factor V G1691A (Leiden) and prothrombin G20210A single nucleotide polymorphisms among Arabs. J Thromb Thrombolysis. 2005 Dec;20(3):163–8. doi: 10.1007/s11239-005-3550-4. PMID: 16261289.

Alvarez-Fernandez, A., Bernal, M. J., Fradejas, I., Martin Ramírez, A., Md Yusuf, N. A., Lanza, M., … & Rubio, J. M. (2021). KASP: a genotyping method to rapid identification of resistance in Plasmodium falciparum. Malaria Journal, 20(1), 1–8.

Al-Zoubi, N., Alrabadi, N., Kheirallah, K., & Alqudah, A. (2021). Prevalence and Multiplicity of Thrombophilia Genetic Polymorphisms of FV, MTHFR, F II, and PAI-I: A Cross-Sectional Study on a Healthy Jordanian Population. International Journal of General Medicine, 5323–5332.

Badireddy M, Mudipalli VR. Deep Venous Thrombosis Prophylaxis. 2022 Aug 22. In: StatPearls [Internet]. Treasure Island (FL): StatPearls Publishing; 2022 Jan–. PMID: 30521286.

Balta, G. Ü. N. A. Y., Altay, C., & Gurgey, A. Y. T. E. M. İ. Z. (2003). Prevalence of PAI-1 gene 4G/5G genotype in Azerbaijan and Kyrgyzstan populations: literature review. Journal of Thrombosis and Haemostasis, 1(4), 858–859.

Bezgin T, Kaymaz C, Akbal Ö, Yılmaz F, Tokgöz HC, Özdemir N. Thrombophilic Gene Mutations in Relation to Different Manifestations of Venous Thromboembolism: A Single Tertiary Center Study. Clin Appl Thromb Hemost. 2018 Jan;24(1):100–106. doi: 10.1177/1076029616672585. Epub 2016 Oct 11. PMID: 27729560; PMCID: PMC6714624.

Dajani, R., Fatahallah, R., Dajani, A., Al-Shboul, M., & Khader, Y. (2012). Prevalence of coagulation factor II G20210A and factor V G1691A Leiden polymorphisms in Chechans, a genetically isolated population in Jordan. Molecular biology reports, 39, 9133–9138.

Dajani, R., Fathallah, R., Arafat, A., AbdulQader, M. E., Hakooz, N., Al-Motassem, Y., & El-Khateeb, M. (2013). Prevalence of MTHFR C677T single nucleotide polymorphism in genetically isolated populations in Jordan. Biochemical genetics, 51, 780–788.

Dean L. Methylenetetrahydrofolate Reductase Deficiency. 2012 Mar 8 [updated 2016 Oct 27]. In: Pratt VM, Scott SA, Pirmohamed M, Esquivel B, Kane MS, Kattman BL, Malheiro AJ, editors. Medical Genetics Summaries [Internet]. Bethesda (MD): National Center for Biotechnology Information (US); 2012–. PMID: 28520345.

Djordjevic V, Stankovic M, Brankovic-Sreckovic V, Rakicevic L, Damnjanovic T, Antonijevic N, Radojkovic D. Prothrombotic genetic risk factors in stroke: a possible different role in pediatric and adult patients. Clin Appl Thromb Hemost. 2012 Nov;18(6):658–61. doi: 10.1177/1076029611432136. Epub 2012 Jan 23. PMID: 22275392.

Dölek, B., Eraslan, S., Eroğlu, S., Kesim, B. E., Ulutin, T., Yalçıner, A., … & Gözükirmızı, N. (2007). Molecular analysis of factor V Leiden, factor V Hong Kong, factor II G20210A, methylenetetrahydrofolate reductase C677T, and A1298C mutations related to Turkish thrombosis patients. Clinical and Applied Thrombosis/Hemostasis, 13(4), 435–438.

Golestani, A., Rahimi, A., Moridi, N. et al. Association of factor V Leiden R506Q, FXIIIVal34Leu, and MTHFR C677T polymorphisms with acute myocardial infarction. Egypt J Med Hum Genet 23, 118 (2022). 10.1186/s43042-022-00330-9.

Gowda, M.S., Zucker, M.L., Vacek, J.L. et al. Incidence of Factor V Leiden in Patients with Acute Myocardial Infarction. J Thromb Thrombolysis 9, 43–45 (2000). 10.1023/A:1018652429633

Graydon, J. S., Claudio, K., Baker, S., Kocherla, M., Ferreira, M., Roche-Lima, A., … & Ruaño, G. (2019). Ethnogeographic prevalence and implications of the 677C> T and 1298A> C MTHFR polymorphisms in US primary care populations. Biomarkers in Medicine, 13(8), 649–661.

He, Chunlin, John Holme, and Jeffrey Anthony. “SNP genotyping: the KASP assay.” Crop breeding. Humana Press, New York, NY, 2014. 75–86.

Hickey, S. E., Curry, C. J., & Toriello, H. V. (2013). ACMG Practice Guideline: lack of evidence for MTHFR polymorphism testing. Genetics in Medicine, 15(2), 153–156.

Isordia-Salas, I., Leaños-Miranda, A., Sainz, I. M., Reyes-Maldonado, E., & Borrayo-Sánchez, G. (2009). Association of the plasminogen activator inhibitor-1 gene 4G/5G polymorphism with ST elevation acute myocardial infarction in young patients. Revista Española de Cardiología (English Edition), 62(4), 365–372.

John A. Heit, 14 - Thrombophilia: Clinical and Laboratory Assessment and Management, Editor(s): Craig S. Kitchens, Craig M. Kessler, Barbara A. Konkle, Consultative Hemostasis and Thrombosis (Third Edition), W.B. Saunders, 2013, Pages 205–239, ISBN 9781455722969, 10.1016/B978-1-4557-2296-9.00014-2. (https://www.sciencedirect.com/science/article/pii/B9781455722969000142)

Joint, W. H. O. (1995). Inherited thrombophilia: report of a Joint WHO (No. WHO/HGN/ISTH/WG/95.5. Unpublished). World Health Organization.

Kazuo Miyashita, Show Nishikawa, Masashi Hosokawa, Chapter 29 - Therapeutic Effect of Fucoxanthin on Metabolic Syndrome and Type 2 Diabetes, Editor(s): Debasis Bagchi, Nair Sreejayan, Nutritional and Therapeutic Interventions for Diabetes and Metabolic Syndrome, Academic Press, 2012, Pages 367–379, ISBN 9780123850836, 10.1016/B978-0-12-385083-6.00029-2.

Kujovich, J. L. (2018). Factor V Leiden thrombophilia.

Linnemann, B., & Hart, C. (2019). Laboratory diagnostics in thrombophilia. Hämostaseologie, 39(01), 049–061.

Madkhaly F, Alshaikh A, Alkhail HA, Alnounou R, Owaidah T. Prevalence of positive factor V Leiden and prothrombin mutations in samples tested for thrombophilia in Saudi Arabia. Am J Blood Res. 2021 Jun 15;11(3):255–260. PMID: 34322288; PMCID: PMC8303010.

Makhoul M, Rambla C, Voss-Fels KP, Hickey LT, Snowdon RJ, Obermeier C. Overcoming polyploidy pitfalls: a user guide for effective SNP conversion into KASP markers in wheat. Theor Appl Genet. 2020 Aug;133(8):2413–2430. doi: 10.1007/s00122-020-03608-x. Epub 2020 Jun 4. PMID: 32500260; PMCID: PMC7360542.

Morange, P. E., Saut, N., Alessi, M. C., Yudkin, J. S., Margaglione, M., Di Minno, G., … & Juhan-Vague, I. (2007). Association of plasminogen activator inhibitor (PAI)-1 (SERPINE1) SNPs with myocardial infarction, plasma PAI-1, and metabolic parameters: the HIFMECH study. Arteriosclerosis, thrombosis, and vascular biology, 27(10), 2250–2257.

Nikolopoulos, G., Bagos, P., Tsangaris, I., Tsiara, C., Kopterides, P., Vaiopoulos, A., Kapsimali, V., Bonovas, S. & Tsantes, A. (2014). The association between plasminogen activator inhibitor type 1 (PAI-1) levels, PAI-1 4G/5G polymorphism, and myocardial infarction: a Mendelian randomization meta-analysis. Clinical Chemistry and Laboratory Medicine (CCLM), 52(7), 937–950. 10.1515/cclm-2013-1124

Özbek, N., Ataç, F. B., Verdi, H., & Kayıran, S. M. (2003). Purpura fulminans in a child with combined heterozygous prothrombin G20210A and factor V Leiden mutations. Annals of hematology, 82, 118–120.

Raptopoulou, A., Michou, V., Mourtzi, N., Papageorgiou, E. G., Voyiatzaki, C., Tsilivakos, V., … & Bei, T. A. (2022). LargeLscale screening for factor V Leiden (G1691A), prothrombin (G20210A), and MTHFR (C677T) mutations in Greek population. Health Science Reports, 5(4), e457.

Sabbagh, A. S., Mahfoud, Z., Taher, A., Zaatari, G., Daher, R., & Mahfouz, R. A. (2008). High prevalence of MTHFR gene A1298C polymorphism in Lebanon. Genetic Testing, 12(1), 75–80.

Settin, A. A., Alghasham, A., Ali, A., Dowaidar, M., & Ismail, H. (2012). Frequency of thrombophilic genetic polymorphisms among Saudi subjects compared with other populations. Hematology, 17(3), 176–182.

Tsai, S. J., Hong, C. J., Liou, Y. J., Yu, Y. W. Y., & Chen, T. J. (2008). Plasminogen activator inhibitor-1 gene is associated with major depression and antidepressant treatment response. Pharmacogenetics and genomics, 18(10), 869–875.

Xu X, Wang H, Li H, Cui X, Zhang H. SERPINE1 −844 and −675 polymorphisms and chronic obstructive pulmonary disease in a Chinese Han population. Journal of International Medical Research. 2016;44(6):1292–1301. doi:10.1177/0300060516664270

Xu, Y., Duan, L., Wong, D. W. K., Wong, T. Y., & Liu, J. (2016). Semantic reconstruction-based nuclear cataract grading from slit-lamp lens images. In Medical Image Computing and Computer-Assisted Intervention-MICCAI 2016: 19th International Conference, Athens, Greece, October 17-21, 2016, Proceedings, Part III 19 (pp. 458–466). Springer International Publishing.

Yang, S., Yu, W., Wei, X., Wang, Z., Zhao, Y., Zhao, X., … & Zhang, X. (2020). An extended KASP-SNP resource for molecular breeding in Chinese cabbage (Brassica rapa L. ssp. pekinensis). Plos one, 15(10), e0240042.

Yapijakis, C., Serefoglou, Z., & Voumvourakis, C. (2015). Common Gene Polymorphisms Associated with Thrombophilia. In (Ed.), Thrombosis, Atherosclerosis and Atherothrombosis - New Insights and Experimental Protocols. IntechOpen. 10.5772/61859

Yapijakis, C., Serefoglou, Z., Nixon, A. M., Vylliotis, A., Ragos, V., & Vairaktaris, E. (2012). Prevalence of thrombosis-related DNA polymorphisms in a healthy Greek population. in vivo, 26(6), 1095–1101.

Yousef, A. M., Shomaf, M., Berger, S., Ababneh, N., Bobali, Y., Ali, D., … & Ismail, S. (2013). Allele and genotype frequencies of the polymorphic methylenetetrahydrofolate reductase and colorectal cancer among Jordanian population. Asian Pacific Journal of Cancer Prevention, 14(8), 4559–4565.

Zhang, Y., Liang, D., Huang, H., Yang, Z., Wang, Y., Yu, Y., … & Xiao, W. (2020). Development and application of KASP assays for rapid screening of 8 genetic defects in Holstein cattle. Journal of Dairy Science, 103(1), 619–624.

